# The Lancet peer reviewers and the COVID-19 pandemic

**DOI:** 10.1101/2023.08.02.23293558

**Authors:** Gwinyai Masukume, Victor Grech

## Abstract

**Introduction:** Peer review is paramount to the scholarly article paradigm, helping to ensure the integrity and credibility of research. *The Lancet* played a crucial role in disseminating key information on the COVID-19 pandemic, publishing early clinical descriptions, risk factors for death, and effectiveness of measures like physical distancing and masks. Notably, *The Lancet* was the world’s most cited journal for COVID-19 research, emphasising its significant impact on disseminating critical findings during the pandemic.

**Methods:** Geographic data for *The Lancet’s* peer reviewers in 2019 (pre-pandemic) and 2020 (pandemic) were analysed at the country level, ranking reviewer countries. A test of proportions compared reviewer numbers between the years.

**Results:** In 2020, China emerged as one of the top ten reviewer countries for the first time, with a significant increase from 1% (25 of 1843) in 2019 to 3% (54 of 1850), p=0.001. Italy also entered the top five reviewer countries, rising from 4% (67) to 5% (90), p=0.065. Reviewers from Africa 43 (2%) and South America 31 (2%) represented their continents in 2020. The top ten reviewer nations for *The Lancet* in 2020 largely mirrored the top ten countries in global COVID-19 research output.

**Conclusion:** During the COVID-19 pandemic’s acute phase in 2020, *The Lancet*, the world’s most cited journal for COVID-19 research, featured peer reviewers who were largely representative of global COVID-19 research output. Notably, reviewers from China, the first country affected by COVID-19, increased significantly. However, underrepresentation of some continents persisted. To foster global idea exchange and enhance pandemic preparedness, research capacity worldwide must expand, broadening the reviewer pool—a vital step given uncertainties in future pandemic geographic origin.

---

*The Lancet* played a crucial role in disseminating key information related to the COVID-19 pandemic which in turn informed global practice. For instance *The Lancet* published early on, in January 2020, one of the first descriptions of the clinical features of this pandemic.^1^ Similarly, *The Lancet* published an early paper, in March 2020, describing the risk factors for death in hospitalised patients.^2^ Indeed *The Lancet* was the world’s most cited journal for COVID-19 research.^3^ These research articles were widely shared and discussed in the news and on social media. An example was a paper that analysed how physical distancing, face masks, and eye protection reduced disease spread.^4^ Another example is a paper that looked into COVID-19 treatment with hydroxychloroquine or chloroquine that was subsequently discovered to be based on false data and hence retracted.^5^

Peer review is critical to the scholarly article paradigm.^6^ It is thus important to explore how peer review changed at *The Lancet* during the acute pandemic phase, in 2020,^7^ since this journal will likely play an equally central role in the publication of impactful findings during future pandemics.

We analysed data for over 3600 of *The Lancet’s* peer reviewers for the years 2019^8^ (pre-pandemic) and 2020^9^ (pandemic). Because the data was publicly available ethics review was not required. A two-sample test of proportions in Stata version 17BE (StataCorp LP College Station, TX) was used to compare the proportion of reviewers between the years. In 2020 China entered the top ten reviewer countries for the first time in comparison to pre-pandemic data (figure 1), while Italy entered the top five reviewer countries for the first time (figure 2).^10^ China increased its number of reviewers from 1% (25 of 1843) in 2019 to 3% (54 of 1850) in 2020, p=0.001. For Italy, the numbers were 4% (67) and 5% (90) respectively, p=0.065. These findings were consistent with the global trajectory of the severe acute respiratory syndrome coronavirus 2 (SARS-CoV-2), with an early peak in COVID-19 papers from China and a subsequent rise in papers from Italy as the virus spread.^11^

**Figure 1:**
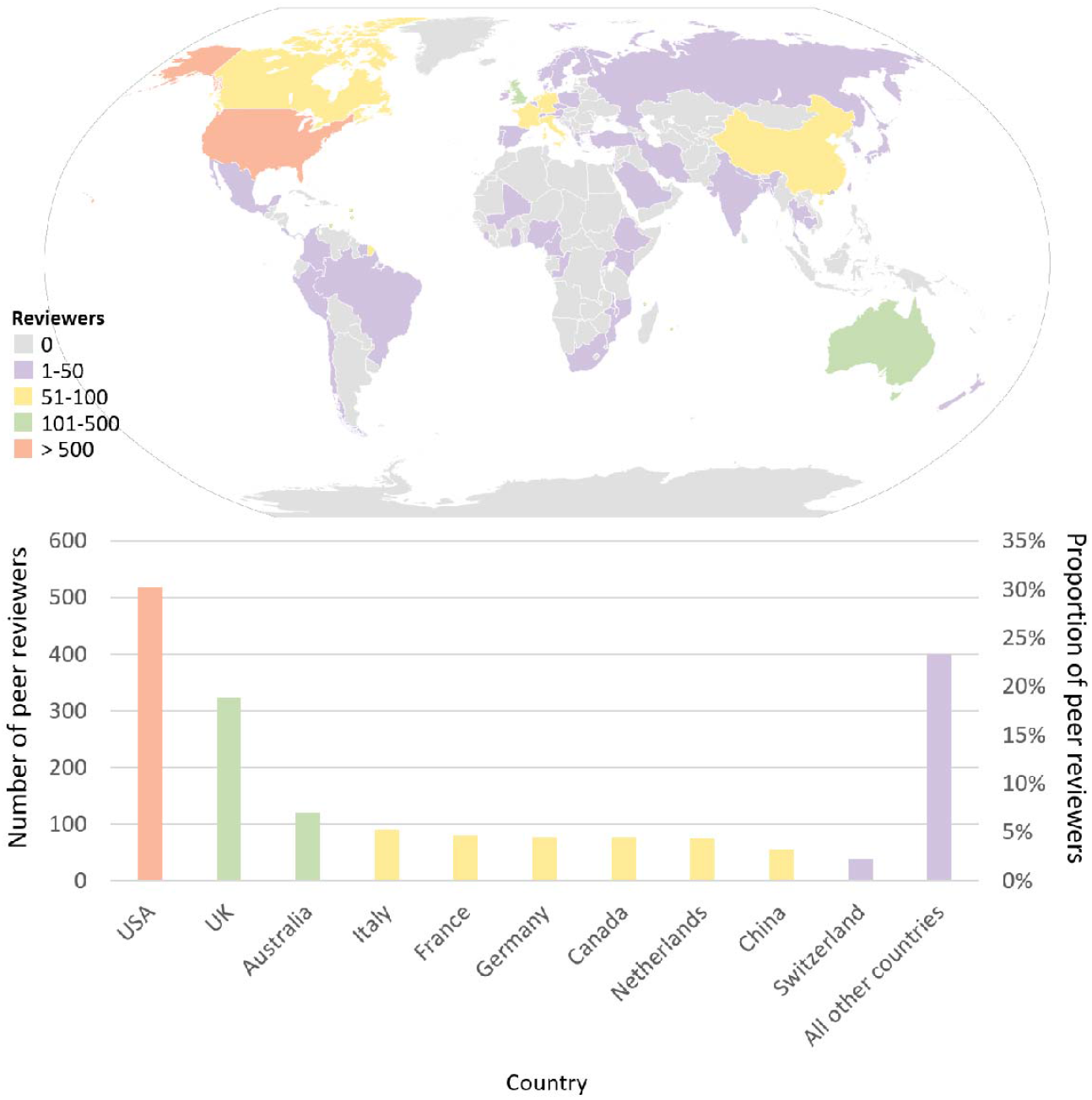
Number and location of *The Lancet* peer reviewers, 2020.

**Figure 2:**
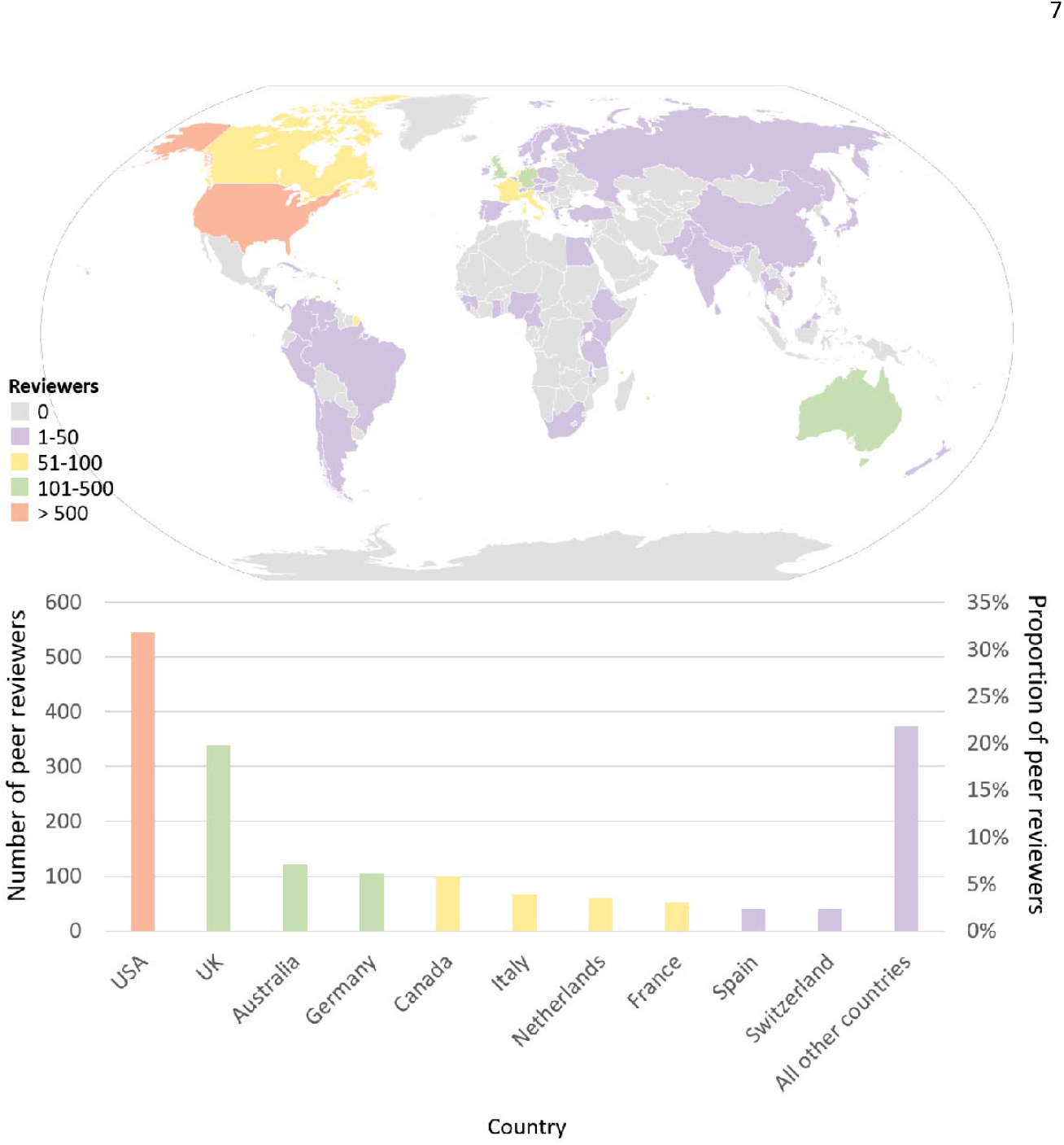
Number and location of *The Lancet* peer reviewers, 2019.

The top two reviewer countries were the USA and the UK which accounted for 48% (884) of reviewers in 2019 and 45% (841) in 2020, p=0.13. This was consistent with the earlier pre-pandemic pattern.^10^ With the exception of China in 2020, the top ten nations in terms of the number of peer reviewers remained the same. These included France, Germany, and Canada, all of which are members of the Group of Seven (G7), the largest advanced economies, along with the aforementioned USA, UK, and Italy.

With two exceptions, the top ten nations by peer reviewer country of origin for *The Lancet* in 2020 were identical to the top ten countries by global COVID-19 research output in 2020.^12^ These two nations were Spain and India.

Reviewers from Africa in 2020 were 43 (2%), 19 of them from South Africa, and there were 31 (2%) from South America, 23 of them from Brazil. These data were in line with pre-pandemic reviewer data.

In conclusion, with a notable increase from China, the first country to be affected by COVID-19, *The Lancet* peer reviewers during the acute phase of the COVID-19 pandemic were largely representative of the global COVID-19 research output. Reviewer numbers also reflected the current underrepresentation of some continents in global research. Prior to the next pandemic, it might be useful to recruit reviewers from underrepresented global regions in order to foster global idea exchanges and potentially create a more effective response to such crises but this may be hindered by smaller numbers – there may be fewer eligible researchers in these regions so that the current underrepresentation simply reflects this, posing a reviewer recruiting challenge to all journals. It is crucial to expand the capacity of research worldwide and, consequently, the pool of reviewers. As the geographic origin of the next pandemic remains uncertain, a more inclusive approach is essential.

## Data Availability

All data produced in the present work are contained in the manuscript

https://doi.org/10.1016/S0140-6736(20)30412-8

https://doi.org/10.1016/S0140-6736(21)00459-1

## References

1. Huang C, Wang Y, Li X, Ren L, Zhao J, Hu Y, et al. Clinical features of patients infected with 2019 novel coronavirus in Wuhan, China. Lancet. 2020;395(10223):497–506.

2. Zhou F, Yu T, Du R, Fan G, Liu Y, Liu Z, et al. Clinical course and risk factors for mortality of adult inpatients with COVID-19 in Wuhan, China: a retrospective cohort study. Lancet. 2020;395(10229):1054–62.

3. Kambhampati SBS, Vasudeva N, Vaishya R, Patralekh MK. Top 50 cited articles on Covid-19 after the first year of the pandemic: A bibliometric analysis. Diabetes Metab Syndr. 2021;15(4):102140.

4. Chu DK, Akl EA, Duda S, Solo K, Yaacoub S, Schünemann HJ. Physical distancing, face masks, and eye protection to prevent person-to-person transmission of SARS-CoV-2 and COVID-19: a systematic review and meta-analysis. Lancet. 2020;395(10242):1973–87.

5. Mehra MR, Desai SS, Ruschitzka F, Patel AN. RETRACTED: Hydroxychloroquine or chloroquine with or without a macrolide for treatment of COVID-19: a multinational registry analysis. Lancet. 2020.

6. Ben Messaoud K, Schroter S, Richards M, Gayet-Ageron A. Analysis of peer reviewers’ response to invitations by gender and geographical region: cohort study of manuscripts reviewed at 21 biomedical journals before and during covid-19 pandemic. Bmj. 2023;381:e075719.

7. Callaway E, Ledford H, Viglione G, Watson T, Witze A. COVID and 2020: An extraordinary year for science. Nature. 2020;588(7839):550–2.

8. The Editors of The Lancet. Thank you to The Lancet’s reviewers of 2019. The Lancet. 2020;395(10226):769.

9. The Editors of The Lancet. Thank you to The Lancet’s reviewers of 2020. The Lancet. 2021;397(10277):864.

10. Masukume G, Grech V. The Lancet peer reviewers: global pattern and distribution. Lancet. 2018;391(10140):2603–4.

11. Else H. How a torrent of COVID science changed research publishing - in seven charts. Nature. 2020;588(7839):553.

12. Wagner CS, Cai X, Zhang Y, Fry CV. One-year in: COVID-19 research at the international level in CORD-19 data. PLoS One. 2022;17(5):e0261624.

